# The course of COVID-19 in patients with chronic spontaneous urticaria receiving omalizumab treatment

**DOI:** 10.1101/2022.01.29.22270077

**Authors:** Emel Atayik, Gokhan Aytekin

## Abstract

**Introduction:** Although there are case reports and guideline recommendations that states omalizumab can be used in chronic spontaneous urticaria (CSU) patients during SARS-CoV-2 pandemic, there are scarce studies showing the course of Coronavirus disease 2019 (COVID-19) in CSU patients receiving omalizumab.

**Materials and Methods:** A total of 370 patients with chronic urticaria were included in the study between June 2020 and December 31, 2020.

**Results:** Sixty patients (16.2%) became infected with the SARS-CoV-2. The rate of pneumonia and hospitalization were 4.1% and 1.9%. There was no significant difference was determined between the CSU patients with omalizumab treatment and the non-receivers in regard to the rate of SARS-CoV-2 (+) (p: 0.567) and in regard to the rate of SARS-CoV-2 related pneumonia and hospitalization (p: 0.331 and p: 0.690). Gender, duration of CSU, serum IgE levels, omalizumab treatment, and atopy were not found to be associated with an increased risk for SARS-CoV-2 positivity in patients with CSU.

**Conclusion:** Our study shows that the use of omalizumab does not increase the risk of COVID-19 infection, COVID-19-related pneumonia and hospitalizations in CSU patients and supports the views that omalizumab can be used safely in patients with CSU during the COVID-19 pandemic.

## Introduction

SARS-CoV-2 was first reported in the Wuhan city of China and soon after, the virus and hence the disease got spread rapidly to the entire world and the World Health Organization (WHO) in March 2020, has declared it as a global pandemic (1). Since there is still no definitive treatment for coronavirus disease 2019 (COVID-19) and mass immunization is not at the desired rate, it is very important to identify the most vulnerable patients and the effects of the drugs they use on the course of COVID-19 to reduce the mortality and morbidity caused by SARS-CoV-2. Additionally, the effect of chronic diseases and drugs used in treatment of these diseases on the course of COVID-19 continues to be the subject of many studies.

Omalizumab is an anti-IgE monoclonal antibody used for the treatment of chronic spontaneous urticaria (CSU) patients aged 12 years and older who are refractory to standard dose of antihistamines (2). It has also been approved for the treatment of severe asthma patients with indoor allergen sensitivity (3). In addition to its anti-IgE activity, omalizumab, especially in pediatric patients with allergic asthma, ameliorates inadequate antiviral response, reduces the duration of rhinovirus infection and decreases the viral shedding (4). Therefore, it can be arguable that IgE-targeting antibody treatment, omalizumab, may have protective effects against viruses like SARS-CoV-2. Although there are case reports (5-7) and guideline recommendations (8) that states omalizumab can be used in CSU patients during SARS-CoV-2 pandemic, there are scarce studies showing the course of COVID-19 in CSU patients receiving omalizumab (9).

Therefore, the aims of present study were to try to expose the course of SARS-CoV-2 in patients with chronic urticaria, to compare the prevalence of SARS-CoV-2 infection, SARS-CoV-2 related pneumonia, and SARS-CoV-2 related hospitalization in patients with chronic urticaria receiving omalizumab treatment and patients who did not receive omalizumab treatment (non-receivers) and to identify possible risk factor for SARS-CoV-2 positivity in CSU patients.

## Material and Methods

Adult patients with chronic urticaria, who were being followed-up in a tertiary allergy clinic in Konya, located in the central Anatolia, Turkey, between June 2020 and December 31, 2020, were selected for the study. Forty-three patients with irregular treatment and insufficient information in their files were excluded from the study. Three hundred seventy patients who were receiving active-continuous treatment for CSU during the study period were included in the study.

Diagnosis of chronic urticaria was made by the presence of recurrent urticaria, angioedema, or both, for a period of six weeks or longer. Oral antihistamine treatments were given as the first-line treatment to the patients who applied with the complaints of urticaria. In patients who did not respond adequately, the first treatment dose was increased up to four times or a second-generation oral antihistamine from a different group or a leukotriene receptor antagonist was added to the treatment in line with the guideline recommendations (10). Patients who did not benefit from these treatments, omalizumab, 300 mg/4 weeks/ subcutaneously, was applied for 12 weeks. At the end of 12 weeks, the treatment of patients who benefited from omalizumab treatment, the treatment was completed to 24 weeks and then omalizumab treatment was stopped.

The diagnosis of SARS-CoV-2 was made by a positive Polymerase Chain Reaction (PCR) test in patients with consistent clinical presentation for COVID-19 or by consistent computed tomography findings.

Whole blood count was measured using the Abbott Cell Dyn 3700 series (Sheath reagent), and quantitative determination of serum immunoglobulin (Ig) E was made by the Siemens BN II/ BN ProSpec system (using particle-enhanced immunonephelometry).

The study was approved by Karatay University Ethics Committee (Decision number 2020/015)

Statistical analysis was performed with the IBM SPSS Statistics Version 22 software package. Normally distributed parameters were presented as mean ± standard deviation and data that were not normally distributed were expressed as median (interquartile range: minimum– maximum). Descriptive data were presented as frequencies and percentages and compared using a Chi-square test. Comparisons between baseline characteristics were performed by independent Student t, Mann-Whitney rank-sum, Fisher exact or Chi-square tests where appropriate. As a result of these statistical analysis, parameters with p<0.2 between SARS-CoV-2 (+) patients and SARS-CoV-2 (-) patients were subjected to regression analysis. Binomial logistic regression analysis was performed to determine independent predictors for SARS-CoV-2 positivity.

## Results

A total of 370 patients with chronic urticaria were included in the study [Female: 258 (69.7%), Male: 112 (30.3%)]. The mean age was 40.50 years (18 to 86 years). The mean duration of disease was 3 years (0.5-29). One hundred twenty patients (32.4%) have at least one allergen sensitivity and the rest of them were non-allergic. Seventy-nine patients (21.4%) had at least one accompanying comorbidity and the most common comorbidity was hypertension (8.4%). One hundred seventy-nine patients (48.4%) were receiving omalizumab treatment for chronic urticaria. Mean serum IgE level was 87.55 (7-3880) IU/ml, eosinophil count was 130 (0-2290) cells/ml and vitamin D level was 15.17 (2-153) μg/L.

Sixty patients (16.2%) became infected with the SARS-CoV-2 virus during the study period. Fifteen patients (4.1%) had pneumonia due to SARS-CoV-2 and seven of them (1.9%) were hospitalized. Clinical characteristics of the patients are summarized in Table 1.

**Table 1.** Demographic, clinical and laboratory parameters of patients with CSU according to omalizumab treatment

| Parameters | Study population<br>n: 370 | Chronic urticaria<br>patients with<br>omalizumab<br>(n: 179 ) | Chronic urticaria<br>patients with out<br>omalizumab<br>(n: 191) | p |
| --- | --- | --- | --- | --- |
| Age, years | 40.50 (18-86) | 42 (18-83) | 38 (18-86) | 0.201 |
| Gender, Female, n (%) | 258 (69.7) | 118 (65.9) | 140 (73.3) | 0.123 |
| Duration of disease, years | 3 (0.5-29) | 3 (0.5-20) | 3 (0.6-29) | 0.714 |
| Atopy, n (%) | 120 (32.4) | 68 (38) | 52 (27.2) | **0.027** |
| Comorbidities, n % | 79 (21.4) | 45 (25.1) | 34 (17.8) | 0.085 |
| • Hypertension | 22 (5.9) | 15 (8.4) | 7 (3.7) | 0.055 |
| • Type 2 DM | 16 (4.3) | 8 (4.5) | 8 (4.2) | 0.894 |
| • CAD | 13 (3.5) | 6 (3.4) | 7 (3.7) | 0.870 |
| IgE, at diagnosis, IU/ml | 87.55 (7-3880) | 108 (10.8-3880) | 81 (7-2500) | **0.001** |
| Vitamin D, µg/L | 15.17 (2-153) | 15.35 (2-153) | 12.40 (10.1-97) | 0.871 |
| Eosinophil count, cell/ml | 130 (0-2290) | 120 (0-1850) | 140 (1.10-2290) | 0.098 |
| SARS-CoV-2, n (%) | 60 (16.2) | 27 (15.1) | 33 (17.3) | 0.567 |
| Pneumonia, n (%) | 15 (4.1) | 9 (5) | 6 (3.1) | 0.331 |
| Hospitalization, n (%) | 7 (1.9) | 4 (2.2) | 3 (1.5) | 0.690 |

CSU: Chronic spontaneous urticaria, DM: Diabetes mellitus, CAD: Coronary artery disease, Ig: Immunoglobulin, SARS-CoV-2: Severe acute respiratory syndrome coronavirus 2.

We divided the study participants into two group as the chronic urticaria patients on omalizumab treatment and chronic urticaria patients not receiving omalizumab treatment; no significant difference was determined between the groups in terms of age, gender, duration of disease, accompanying comorbidities (Type 2 diabetes mellitus [DM], hypertension [HT], and coronary artery disease [CAD]), baseline eosinophil count and vitamin D level and frequency of infection with SARS-CoV-2 virus, SARS-CoV-2 related pneumonia and SARS-CoV-2 related hospitalization. However, there was a significant difference between the groups in terms of sensitivity to allergens (atopy) and serum IgE levels (p: 0.027 and p: 0.001, respectively).

Within the 370 patients included in the study, the rate of pneumonia was 4.1% (15 patients). Twenty-seven patients (15.1%) in omalizumab group and 33 patients (17.3%) in non omalizumab group became infected with the SARS-CoV-2 virus during the study period. There was no significant difference was determined between the chronic urticaria patients with omalizumab treatment and the non-receivers in regard to the rate of SARS-CoV-2 (+) (p: 0.567).

Nine patients (5%) in omalizumab group and six patients (3.1%) in non-omalizumab group had pneumonia due to SARS-CoV-2. Four patients (2.2%) in omalizumab group and three patients (1.5%) in non-omalizumab group were hospitalized. There was no significant difference was determined between the chronic urticaria patients with omalizumab treatment and the non-receivers in regard to the rate of SARS-CoV-2 realted pneumonia and SARS-CoV-2 related hospitalization (p: 0.331 and p: 0.690) (Table 1). There were no patients admitted to the intensive care unit during the study period. We did not lost any of our patients due to COVID-19 infection.

When SARS-CoV-2 positive and negative CSU patients were compared; there were no significant differences between both groups in terms of gender, current age, duration of disease, accompanying comorbidities, serum IgE levels, vitamin D levels and rate of patients receiving omalizumab therapy. A significant difference was determined in terms of eosinophil count (p: 0.015) (Table 2).

**Table 2.** Demographic and clinical characteristics of patients with CSU according to SARS-CoV-2.

| Parameters | SARS-CoV-2 (+)<br>(n: 60) | SARS-CoV-2 (-)<br>(n: 310) | p |
| --- | --- | --- | --- |
| Age, years | 45.5 (19-83) | 41.50 (18-83) | 0.453 |
| Gender, Female, n (%) | 40 (66.7) | 218 (70.3) | 0.573 |
| Duration of disease, years | 2.5 (0.6-20) | 3 (0.5-29) | 0.417 |
| Atopy, n (%) | 17 (28.3)) | 103 (33.2) | 0.459 |
| Omalizumab treatment, n (%) | 27 (45) | 152 (49.0) | 0.567 |
| Comorbidities, n (%) | 12 (20) | 67 (21.6) | 0.780 |
| • Hypertension | 4 (6.7) | 12 (3.9) | 0.330 |
| • Type 2 DM | 3 (5) | 19 (6.1) | 0.735 |
| • CAD | 2 (3.3) | 11 (3.5) | 0.934 |
| IgE, at diagnosis, IU/ml | 103 (16.70-2580) | 119 (10-3880) | 0.876 |
| Eosinophil count, cell/ml | 160 (0-1850) | 120 (0-1390) | **0.015** |
| Vitamin D, µg/L | 15.14 (6.3-45) | 15.23 (2-153) | 0.988 |

CSU: Chronic spontaneous urticaria, SARS-CoV-2: Severe acute respiratory syndrome coronavirus 2, DM: Diabetes mellitus, CAD: Coronary artery disease, Ig: Immunoglobulin.

Univariant and multivariant binary logistic regression analyses showed that, current age, gender, duration of disease, the accompanying comorbidities, eosinophil count, serum IgE levels, omalizumab treatment, and atopy were not found to be associated with an increased risk for SARS-CoV-2 positivity in patients with CSU (Table 3).

**Table 3.** Logistic regression analysis of possible risk factors associated with SARS-CoV-2 in patients with CSU

| Variables | Univariate Analysis |  | Multivariate Analysis |  |
| --- | --- | --- | --- | --- |
|  | OR (95% CI) | P-value | OR (95% CI) | P-value |
| Age | 1.009 (0.991-1.028) | 0.314 | 1.012 (0.992-1.032) | 0.260 |
| Gender | 0.844 (0.468-1.522) | 0.573 | 0.887 (0.480-1.640) | 0.703 |
| Duration of disease | 0.997 (0.932-1.067) | 0.933 | 0.980 (0.913-1.052) | 0.575 |
| Comorbidities | 0.907 (0.456-1.804) | 0.780 | 0.830 (0.390-1.766) | 0.629 |
| Eosinophil count | 1.001 (1.000-1.002) | 0.120 | 1.001 (1.000-1.002) | 0.156 |
| IgE | 1.000 (1.000-1.001) | 0.718 | 1.000 (0.999-1.001) | 0.883 |
| Omalizumab treatment | 0.850 (0.488-1.492) | 0.568 | 0.844 (0.474-1.504) | 0.565 |
| Atopy | 0.795 (0.432-0.795) | 0.459 | 0.665 (0.331-1334) | 0.250 |

| Variables | Univariate Analysis |  | Multivariate Analysis |  |
| --- | --- | --- | --- | --- |
|  | OR (95% CI) | P-value | OR (95% CI) | P-value |
| Age | 1.009 (0.991-1.028) | 0.314 | 1.012 (0.992-1.032) | 0.260 |
| Gender | 0.844 (0.468-1.522) | 0.573 | 0.887 (0.480-1.640) | 0.703 |
| Duration of disease | 0.997 (0.932-1.067) | 0.933 | 0.980 (0.913-1.052) | 0.575 |
| Comorbidities | 0.907 (0.456-1.804) | 0.780 | 0.830 (0.390-1766) | 0.629 |
| Eosinophil count | 1.001 (1.000-1.002) | 0.120 | 1.001 (1.000-1.002) | 0.156 |
| IgE | 1.000 (1.000-1.001) | 0.718 | 1.000 (0.999-1.001) | 0.883 |
| Omalizumab treatment | 0.850 (0.488-1.492) | 0.568 | 0.844 (0.474-1504) | 0.565 |
| Atopy | 0.795 (0.432-0.795) | 0.459 | 0.665 (0.331-1334) | 0.250 |

SARS-CoV-2: Severe acute respiratory syndrome coronavirus 2, CSU: Chronic spontaneous urticaria, Ig: Immunoglobulin.

## Discussion

The presented study had three clinically important findings. Firstly, patients with chronic urticaria receiving omalizumab had higher atopy rates and serum IgE values than patients with chronic urticaria who did not receive omalizumab. Secondly, the prevalence of SARS-CoV-2, SARS-CoV-2-associated pneumonia, and SARS-CoV-2-related hospitalization rates are similar in chronic urticaria patients receiving omalizumab and chronic urticaria patients not receiving omalizumab. Lastly, age, gender, presence of atopy, duration of CSU or use of omalizumab are not a risk factor for SARS-CoV-2 positivity (+) in patients with chronic urticaria.

In our study, atopy rates and serum IgE values were higher in chronic urticaria patients who received omalizumab than in chronic urticaria patients who did not receive omalizumab. Almost 50% of CSU patients have elevated IgE levels. It has been demonstrated that elevated IgE levels in CSU patients are associated with increased disease activity, longer disease duration and rapid response to omalizumab (11). Given that the Omalizumab is second line treatment for patients who have inadequate response to antihistamine treatments, elevated IgE levels in patients receiving omalizumab are not surprising. Allergic skin diseases like atopic dermatitis and urticaria are closely associated comorbidities with allergic rhinitis and rhinosinusitis. Chronic urticaria is the most common comorbid disease in patients with atopic dermatitis and it has been shown that the risk of CSU is 2.5 times higher in patients with atopic dermatitis (12). High rates of co-existence of allergic diseases appears to be associated with common and/or similar pathological pathways. For this reason, we think that CSU patients receiving omalizumab have a higher rate of atopy.

In our study, we found that prevalence of SARS-CoV-2, SARS-CoV-2 associated pneumonia and SARS-CoV-2 related hospitalization rates are similar in patients with chronic urticaria who received omalizumab or did not receive omalizumab. Abduelmula et al. reported that none of the 184 CSU and asthma patients who received omalizumab were tested positive for SARS-CoV-2 (13). In another study, Bostan et al. reported that 15 (9.6%) of the 233 patients with CSU were tested positive for SARS-CoV-2 and 2 patients were hospitalized due to SARS-CoV-2 disease (14). Kocaturk et al. also reported that 11 (13.9%) of the 79 patients with CSU who had SARS-CoV-2 positivity 11 patients (13.9%) were hospitalized (8 mild disease and 3 severe disease) (15). In our study, the prevalence of SARS-CoV-2 was 16.2% and the hospitalization rate was 1.9%, which is similar to the aforementioned studies.

Ayhan et al. reported 3 patients with COVID-19 who were receiving omalizumab, one patient was hospitalized for SARS-CoV-2-related pneumonia, one patient was followed at home due to SARS-CoV-2-related mild pneumonia without being hospitalized, and lung involvement did not develop in the last patient. All three patients recovered from COVID-19 with relatively mild disease (16). Kocaturk et al. reported that of the 70 COVID 19 (+) CSU patients (42 patients on antihistamine, 26 patients on omalizumab and 2 patients on cyclosporine treatment) 6 patients receiving antihistamine and 3 patients receiving omalizumab were hospitalized and they found no relation between CSU treatment and the prevalence of COVID-19 (15). In our study, no patient was followed up in the intensive care unit and no mortality was observed. These results support the idea that patients with CSU are not at high risk for severe COVID-19 (15).

While a fast and well-coordinated innate immune response constitutes the first and most important step of defense against viral infections; dysregulated and excessive immune response can cause an immune damage that will harm the human body (17). In SARS-CoV-2 infection, high virus titers and dysregulation of cytokine/chemokine response - these inflammatory cytokines activate T helper type 1 (Th1) cell response - cause inflammatory cytokine storm and ARDS (acute respiratory distress syndrome) caused by cytokine storm is the leading cause of death in the patients infected with SARS-CoV-2 (18). Theoretically, it has been thought that treatments targeting type 1 inflammation might be beneficial especially in the advanced stages of SARS-CoV-2 where Th1 response is dominant, and therefore, a wide range of treatments such as corticosteroids, interleukin (IL)-1 family antagonists, IL-6 antagonist, TNF blockers, intravenous immunoglobulin have been used in the cases of COVID-19 throughout the pandemic (19). Omalizumab is a recombinant anti-IgE antibody used in the treatment of H1 antihistamine-resistant CSU. In addition to its anti-IgE properties, omalizumab has also been shown to exhibit antiviral effects (20). Omalizumab, especially in pediatric patients with allergic asthma, improves inadequate antiviral response, reduces duration of rhinovirus infection time and viral shedding (15). While decreasing plasmacytoid dendritic cell (pDC) FceRIα protein expression, omalizumab simultaneously increase IFN-α response to rhinovirus and influenza infections. Furthermore, in an experimental study, intramuscular injection of small peptide segment of the omalizumab was shown to reduce cytokines like IL-6, IL-1β and tumor necrosis factor (TNF)-α, which are also responsible for the cytokine storm caused by COVID-19 (21). Unexpectedly, omalizumab has also been shown to negatively regulate innate immunity by decreasing toll-like receptor 7 expression, an increase of which provides an effective IFN-α response (22).

To best our knowledge, there is only one study in the literature that examines the risk factors for SARS-CoV-2 positive disease in CSU patients. In this study, it has been reported that gender, comorbid systemic diseases, CSU treatments (omalizumab and/or antihistamine treatments) were not identified as risk factors for SARS-CoV-2 positivity in CSU patients (14). In our study, result of regression analysis showed that demographic parameters such as age, gender, omalizumab treatment and laboratory parameters such as IgE levels, eosinophil counts were not predictive of SARS-CoV-2 positivity.

The main limitations of our study are its retrospective design and inclusion of data from single center. Furthermore, especially at the beginning of pandemic, given the fact that rapid tests and PCR tests might not be used sufficiently to detect SARS-CoV-2, this may have caused a lower prevalence of SARS-CoV-2. The age population of our study was also younger according to older age groups who are more vulnerable to COVID-19-related morbidity and mortality.

In conclusion, our study shows that the use of omalizumab does not increase the risk of COVID-19 infection, COVID-19-related pneumonia and COVID-19-related hospitalizations in CSU patients and supports the views that omalizumab can be used safely in patients with CSU during the COVID-19 pandemic.

## Data Availability

All data produced in the present study are available upon reasonable request to the authors

## Notes

### Competing Interest Statement

The authors have declared no competing interest.

### Funding Statement

This study did not receive any funding

### Author Declarations

The study was approved by Karatay University Ethics Committee (Decision number 2020/015)

## References

1. Jin Y, Yang H, Ji W, Wu W, Chen S, Zhang W, et al. Virology, Epidemiology, Pathogenesis, and Control of COVID-19. Viruses. 2020;12(4).

2. Passante M, Napolitano M, Dastoli S, Bennardo L, Fabbrocini G, Nistico SP, et al. Safety of omalizumab treatment in patients with chronic spontaneous urticaria and COVID-19. Dermatol Ther. 2021.

3. Bousquet J, Wenzel S, Holgate S, Lumry W, Freeman P, Fox H. Predicting response to omalizumab, an anti-IgE antibody, in patients with allergic asthma. Chest. 2004;125(4):1378–86.

4. Abdelmaksoud A, Goldust M, Vestita M. Omalizumab and COVID-19 treatment: Could it help? Dermatol Ther. 2020;33(4):e13792.

5. Abdelmaksoud A, Goldust M, Vestita M. Comment on “ Chronic spontaneous urticaria exacerbation in a patient with COVID-19: rapid and excellent response to omalizumab”. Int J Dermatol. 2020;59(11):1417–8.

6. Ayhan E, Öztürk M, An İ, Bekçİbaşi M. COVID-19 infection under omalizumab therapy for chronic spontaneous urticaria: three cases. Int J Dermatol. 2021;60(2):253–4.

7. Passante M, Napolitano M, Dastoli S, Bennardo L, Fabbrocini G, Nisticò SP, et al. Safety of omalizumab treatment in patients with chronic spontaneous urticaria and COVID-19. Dermatol Ther. 2021.

8. Klimek L, Pfaar O, Worm M, Eiwegger T, Hagemann J, Ollert M, et al. Use of biologicals in allergic and type-2 inflammatory diseases during the current COVID-19 pandemic: Position paper of Ärzteverband Deutscher Allergologen (AeDA)(A), Deutsche Gesellschaft für Allergologie und Klinische Immunologie (DGAKI)(B), Gesellschaft für Pädiatrische Allergologie und Umweltmedizin (GPA)(C), Österreichische Gesellschaft für Allergologie und Immunologie (ÖGAI)(D), Luxemburgische Gesellschaft für Allergologie und Immunologie (LGAI)(E), Österreichische Gesellschaft für Pneumologie (ÖGP)(F) in cooperation with the German, Austrian, and Swiss ARIA groups(G), and the European Academy of Allergy and Clinical Immunology (EAACI)(H). Allergol Select. 2020;4:53–68.

9. Klimek L, Pfaar O, Worm M, Eiwegger T, Hagemann J, Ollert M, et al. Use of biologicals in allergic and type-2 inflammatory diseases during the current COVID-19 pandemic: Position paper of Arzteverband Deutscher Allergologen (AeDA)(A), Deutsche Gesellschaft fur Allergologie und Klinische Immunologie (DGAKI)(B), Gesellschaft fur Padiatrische Allergologie und Umweltmedizin (GPA)(C), Osterreichische Gesellschaft fur Allergologie und Immunologie (OGAI)(D), Luxemburgische Gesellschaft fur Allergologie und Immunologie (LGAI)(E), Osterreichische Gesellschaft fur Pneumologie (OGP)(F) in cooperation with the German, Austrian, and Swiss ARIA groups(G), and the European Academy of Allergy and Clinical Immunology (EAACI)(H). Allergol Select. 2020;4:53–68.

10. Zuberbier T, Aberer W, Asero R, Abdul Latiff AH, Baker D, Ballmer-Weber B, et al. The EAACI/GA(2)LEN/EDF/WAO guideline for the definition, classification, diagnosis and management of urticaria. Allergy. 2018;73(7):1393–414.

11. Altrichter S, Fok JS, Jiao Q, Kolkhir P, Pyatilova P, Romero SM, et al. Total IgE as a Marker for Chronic Spontaneous Urticaria. Allergy Asthma Immunol Res. 2021;13(2):206–18.

12. Darlenski R, Kazandjieva J, Zuberbier T, Tsankov N. Chronic urticaria as a systemic disease. Clin Dermatol. 2014;32(3):420–3.

13. Abduelmula A, Georgakopoulos JR, Mufti A, Devani AR, Gooderham MJ, Hong CH, et al. Incidence of COVID-19 in Patients With Chronic Idiopathic Urticaria and Asthma on Omalizumab: A Multicentre Retrospective Cohort Study. Journal of cutaneous medicine and surgery. 2021:12034754211049707.

14. Bostan E, Zaid F, Karaduman A, Dogan S, Gulseren D, Yalici-Armagan B, et al. The effect of COVID-19 on patients with chronic spontaneous urticaria treated with omalizumab and antihistamines: A cross-sectional, comparative study. Journal of cosmetic dermatology. 2021;20(11):3369–75.

15. Kocaturk E, Salman A, Cherrez-Ojeda I, Criado PR, Peter J, Comert-Ozer E, et al. The global impact of the COVID-19 pandemic on the management and course of chronic urticaria. Allergy. 2021;76(3):816–30.

16. Ayhan E, Ozturk M, An I, BekcIbasi M. COVID-19 infection under omalizumab therapy for chronic spontaneous urticaria: three cases. Int J Dermatol. 2021;60(2):253–4.

17. Soy M, Keser G, Atagunduz P, Tabak F, Atagunduz I, Kayhan S. Cytokine storm in COVID-19: pathogenesis and overview of anti-inflammatory agents used in treatment. Clin Rheumatol. 2020;39(7):2085–94.

18. Hu B, Huang S, Yin L. The cytokine storm and COVID-19. J Med Virol. 2021;93(1):250–6.

19. Ye Q, Wang B, Mao J. The pathogenesis and treatment of the ‘Cytokine Storm’ in COVID-19. J Infect. 2020;80(6):607–13.

20. Phipatanakul W, Mauger DT, Guilbert TW, Bacharier LB, Durrani S, Jackson DJ, et al. Preventing asthma in high risk kids (PARK) with omalizumab: Design, rationale, methods, lessons learned and adaptation. Contemp Clin Trials. 2021;100:106228.

21. Wang T, Hou W, Fu Z. Preventative effect of OMZ-SPT on lipopolysaccharide-induced acute lung injury and inflammation via nuclear factor-kappa B signaling in mice. Biochem Biophys Res Commun. 2017;485(2):284–9.

22. Cardet JC, Casale TB. New insights into the utility of omalizumab. J Allergy Clin Immunol. 2019;143(3):923–6 e1.

